# Psychosocial Stressors and Psychiatric Morbidity Among Afghan Refugees Attending a Psychiatry Clinic in Islamabad, Pakistan: A Descriptive Cross-Sectional Study

**DOI:** 10.64898/2026.09.03.26362105

**Authors:** Mahrukh Nadeem, Ahmed Ali Khan, Muhammad Hussain Mansoor, Abdul Wahab Yousafzai

## Abstract

**BACKGROUND:** War refugees are prone to psychiatric ailments due to multiple psychosocial stressors. Afghan refugees remain underrepresented in psychiatric studies. Therefore, a study exploring factors contributing to their mental health disorders is essential to guide rehabilitation efforts worldwide.

**METHODS:** This descriptive cross-sectional study evaluates the demographics, psychosocial stressors, psychiatric illnesses, and psychotropic drug use among Afghan refugees visiting the Psychiatry Outpatient Department (OPD) in Islamabad, Pakistan. Convenience sampling was employed on a sample population (n) of 75 participants. Data was collected via a structured questionnaire and analyzed using SPSS v21.0. Qualitative and quantitative variables were analyzed statistically.

**RESULTS:** Nearly 64%(n=48) of the participants were female. About 46.7%(n=35) of participants were uneducated, and 53.3%(n=40) were unemployed. 86.7%(n=65) of participants were married, while 2.7%(n=2) were divorced. 91%(n=61) of the married and divorced participants had an arranged marriage, 46.3%(n=31) had 5-8 children, and 67.2%(n=45) had experienced physical and/or emotional abuse. 74.7%(n=56) of the total participants reported a loss in the family, while 77.3%(n=58) had experienced trauma. 62.7%(n=47) had depression and associated psychotropic use, e.g., SSRIs (42.7%), TCAs (16%), and Benzodiazepines (13.3%). Alarmingly, 37.3%(n=28) reported a previous suicide attempt.

**CONCLUSION:** This study explores the psychosocial stressors faced by Afghan refugees in Pakistan, including displacement, abuse, trauma, and family/income loss. It emphasizes the importance of evidence-based psychiatric care, rehabilitative interventions, and culturally sensitive care to counter psychiatric morbidities and underscores the need for future research to improve the mental health of refugees globally.

## INTRODUCTION

Afghanistan, a multi-ethnic country with Pashto and Dari as its official languages [1], has endured decades of conflict, disrupting its economy and infrastructure. The war in Afghanistan has left many Afghans internally displaced and forced to seek refuge in neighboring countries. This displacement has profoundly affected national well-being and social cohesion by fragmenting communities and families and adversely affecting the physical and mental health of adults and children [2].

Mental health among Afghan adults is considered a major public health concern [3]. According to the World Health Organization, an estimated 1 million Afghans were suffering from depressive disorders, while 1.2 million were affected by anxiety disorders [4]. Although war-related trauma is significant, cultural and economic factors also contribute to mental health problems [5]. In a study on Afghan adults, war exposure explained only 15% of PTSD symptoms, but adding daily stressors such as poverty, overcrowded housing, unemployment, insecurity, domestic violence, poor health, air pollution, and traffic congestion significantly improved the model’s explanatory power, highlighting the role of both trauma and ongoing hardships in shaping mental health (Miller & Rasmussen, 2010) [6]. Similarly, Miller et al. (2008) found that among women, both war-related experiences and daily stressors predicted depression and PTSD, while in men, only daily stressors were significant predictors [7]. In addition, living in Kabul was identified as an important risk factor [8].

Nonetheless, the impact of war on the mental health of Afghans remains widespread [9]. In a study conducted in Kandahar province of Afghanistan, nearly 60% of parents who lost a child to war had also lost other close relatives, reflecting the widespread familial impact of decades of armed violence in Afghanistan [10]. Furthermore, the convenient access to low-cost illicit drugs has resulted in widespread drug abuse. According to estimates from the United Nations Office on Drugs and Crime (UNODC), about one million Afghans are drug users [11]. Decades of conflict and widespread drug abuse have fostered daily stressors that fuel violence by male relatives, often targeting not only wives but also mothers, sisters, and daughters [12].

Several studies have reported a high burden of psychiatric illnesses among women, which are attributed to sociocultural factors such as early marriage and limited autonomy [13]. In a study involving 160 Afghan women from Kabul and Pakistan during the Taliban regime, 97% suffered from depression, 86% reported anxiety, 42% showed signs of post-traumatic stress disorder (PTSD), and 84% had lost one or more family members [14].

According to the United Nations High Commissioner for Refugees’ Operational Data Portal (ODP) for Afghan Refugees in Pakistan, Pakistan hosts approximately 1.36 million registered Afghan refugees as of February 2025. Moreover, it is estimated that Pakistan received an additional unregistered 600,000 refugees after 2021 [15]. Most of this population resides in Khyber Pakhtunkhwa (KPK) province and therefore, were displaced again during the anti-terrorism military operations in that region. The challenging landscape, underdeveloped infrastructure, deteriorating law-and-order situation, and an under-funded healthcare system make health surveys inadequate in assessing the physical and mental health of this refugee demographic. The concerning psychiatric illness indices of the population of origin, along with the stresses of migration, fragmented family structures, loss of community support, socio-economic hardship, and difficulty adjusting to life in the host country, render this specific population high-risk for developing psychiatric ailments and necessitate further insight into the psycho-social stressors of the Afghan refugees to determine their mental well-being.

Despite the health concerns and shared cultural trauma and resettlement stressors, data on the mental health indices of Afghan refugees residing in Pakistan are limited. This gap is particularly concerning, as refugees are known to experience disproportionately higher rates of both physical and psychological distress yet face significant barriers to healthcare access due to a lack of proper documentation, contributing to widening health inequalities in host countries [16]. With around 43,000 registered Afghan refugees living in Islamabad and given the city’s proximity to the largest refugee population in the underdeveloped Khyber Pakhtunkhwa (KPK) province, this Psychiatry Outpatient Department (OPD) in Islamabad offers a valuable setting to understand their mental health needs. Given the lack of existing surveillance data and the need to identify patterns of distress among refugee populations, a descriptive cross-sectional approach is appropriate to characterize the current mental health status of Afghan refugees. This study explores the demographic profile, psychosocial stressors, and psychiatric conditions of Afghan patients attending the Psychiatry OPD, aiming to identify high-risk individuals and guide interventions to improve the well-being of Afghan refugees in Pakistan and other countries.

## METHODS

This descriptive cross-sectional study was conducted among Afghan refugee patients visiting the Psychiatry OPD in Islamabad, Pakistan, from June 2022 to December 2022. All eligible refugees who visited the OPD during the study period were approached. Seventy-five patients consented and were therefore included in our study. Diagnoses such as Major Depressive Disorder, Obsessive-Compulsive Disorder, Bipolar Disorder, and other mental health disorders were made by qualified psychiatrists in the outpatient clinic based on clinical interviews and assessment using the Diagnostic and Statistical Manual of Mental Disorders, Fifth Edition (DSM-5) criteria. These diagnoses were recorded in patients’ medical records and were eventually used in the study with appropriate confidentiality and ethical considerations.

Patients who spoke Pashto or Persian were included and were communicated with in their native language to ensure inclusivity. There were 1.44 million Afghan refugees as of 30 April 2021; only 2.4% of these refugees resided in Islamabad. Only 57% of this population subset is above 18 years of age [17]. Given the specific focus of this study on Afghan refugees with psychiatric conditions seeking care at a psychiatry clinic in Islamabad, convenience sampling was used to ensure accessibility and feasibility. Similar studies in the past have utilized convenience sampling successfully [18,19]. Patients under the age of 18 and patients with pre-existing psychotic disorders, dementia, debilitating illnesses, physical disabilities, and learning disabilities were excluded from the study.

The data was collected using a self-structured questionnaire (Supplementary Appendix 1 & Supplementary Appendix 2), specifically designed by the consulting Psychiatrist to capture demographic information and psychiatric history relevant to the study objectives. The decision to use a self-structured tool allowed the researchers to tailor questions to the cultural and contextual needs of Afghan refugee patients, which are often not adequately addressed in standardized instruments. After taking informed consent (Supplementary Appendix 2), the questionnaire was administered in an interviewer-led format, where trained researchers filled out the responses after verbally asking each question to the patients in their preferred language. This approach was adopted to accommodate participants with varying literacy levels and to ensure clarity and consistency in understanding the questions. Data was kept under lock and key, and all records were kept confidential to ensure patient confidentiality. Soft copies were password-protected, with only the researchers having access to them.

Academic databases such as PubMed, Google Scholar, and Scopus were used to search for the literature. The search strategy employed Boolean operators to refine results, using combinations of keywords such as *“Afghan refugees” AND “mental health”*, *“depression” OR “bipolar disorder” OR “OCD”*, and *“Pakistan” AND “psychiatric disorders”*. For demographic data, UNHCR (United Nations High Commissioner for Refugees) databases were explored. Data analysis was descriptive and conducted using IBM SPSS Statistics for Windows, Version 21.0. Frequencies and percentages were calculated through the “Frequencies” procedure, and cross-tabulations were performed to examine the relationship between gender and education level. No inferential tests were performed due to the study’s exploratory objectives and limited sample size. The dataset was checked for completeness before analysis. No missing values were identified for the variables included in the reported analyses; therefore, all percentages were calculated using the applicable complete denominator.

## RESULTS

A total of 75 respondents participated in the study. Of the total respondents, 64% (n = 48) were female, while the remaining 36% (n = 27) were male. The mean age of the respondents was 39.37 (SD ±12.10 years), with minimum and maximum ages being 18 and 66 years, respectively. Regarding education, 1.33% (n = 1) of participants had only primary-level education, 42.67% (n = 32) had Secondary School Certificate (SSC), Higher Secondary School Certificate (HSSC), and undergraduate-level education. Another 9.33% (n = 7) of participants had a postgraduate level of education and a diploma, while a majority of 46.67% (n = 35) of participants had no formal education.

Most of the participants were married, 86.7% (n = 65), while 10.7% (n = 8) were unmarried, and 2.7% (n = 2) were divorced. Among the married participants (n = 65), the majority of 90.8% (n = 59) had an arranged marriage, while 9.2% (n = 6) had a love marriage, and 7.7% (n = 5) were in polygamous relationships. The divorced participants (n = 2) were women, both of whom had arranged marriages and monogamous relationships. Among all married and divorced participants (n = 67), 91% (n = 61) had arranged marriages, while 9% (n = 6) had love marriages. Additionally, 7.5% (n = 5) of this group were in polygamous relationships, including three married women whose husbands had other wives and two married men who had multiple wives.

According to the married and divorced participants’ responses, 3% (n = 2) had no children, 35.8% (n = 24) had 1-4 children, 46.3% (n = 31) had 5-8 children, and 14.9% (n = 10) had more than 8 children, with the maximum number of children being 12. Among the respondents, 53.3% (n = 40) were unemployed, 38.7% (n = 29) were working, and 8% (n = 6) were students. [Table 1]

**Table 1:** Sociodemographic, and academic characteristics of the respondents.

| Variable | Frequency (n) | Percentage (%) |
| --- | --- | --- |
| **Gender** |  |  |
| Male | 27 | 36 |
| Female | 48 | 64 |
| **Age** |  |  |
| 18–25 | 12 | 16 |
| 26–35 | 19 | 25.3 |
| 36–45 | 23 | 30.7 |
| 46–55 | 14 | 18.7 |
| 56–66 | 7 | 9.3 |
| **Education** |  |  |
| Uneducated | 35 | 46.7 |
| Primary | 1 | 1.3 |
| SSC & HSSC & Undergraduate | 32 | 42.7 |
| Graduation & Diploma | 7 | 9.3 |
| **Marital status** |  |  |
| Single | 8 | 10.7 |
| Married | 65 | 86.7 |
| Divorced | 2 | 2.7 |
| **Marriage Type\*** |  |  |
| Love | 6 | 9 |
| Arranged | 61 | 91 |
| **Polygamy\*,†** |  |  |
| Yes | 5 | 7.5 |
| No | 62 | 92.5 |
| **Children\*,‡** |  |  |
| No Children | 2 | 3 |
| 1–4 | 24 | 35.8 |
| 5–8 | 31 | 46.3 |
| More than 8 | 10 | 14.9 |
| **Occupation** |  |  |
| Working | 29 | 38.7 |
| Unemployed | 40 | 53.3 |
| Student | 6 | 8 |
| *Percentages for marriage type, polygamy, and children were calculated only for ever-married participants (65 married, 2 divorced). |  |  |
| †Unmarried participants (n = 8) were not in monogamous or polygamous relationships. |  |  |
| ‡Unmarried participants (n = 8) did not have any children. |  |  |

Among the male participants (n = 27), 85.2% (n = 23) were educated, while 14.8% (n = 4) had no formal education. In contrast, among the female participants (n = 48), only 35.4% (n = 17) were educated, whereas a notably higher proportion of 64.6% (n = 31) were uneducated. These descriptive findings demonstrate a marked difference in educational attainment between male and female participants, with a lack of formal schooling being prevalent among the female refugee population. [Table 2]

**Table 2:** Distribution of education status among male and female Afghan refugees.

| Gender | Education Status | Frequency (n) | Percentage (%) within gender groups* |
| --- | --- | --- | --- |
| **Male** | Educated | 23 | 85.2 |
|  | Uneducated | 4 | 14.8 |
| **Female** | Educated | 17 | 35.4 |
|  | Uneducated | 31 | 64.6 |
| **Total** | Educated | 40 | 53.3 |
|  | Uneducated | 35 | 46.7 |

\*Percentages are calculated separately for each gender group based on their respective totals (n = 27 males, n = 48 females).

This study revealed that 69.2% (n = 45) of the married respondents had a history of abuse. Among the abused respondents, a significant portion, 40% (n = 18), were victims of both emotional and physical abuse, while 35.6% (n = 16) faced physical abuse, and 24.4% (n = 11) faced emotional abuse only. Divorced respondents (n = 2) reported no abuse history.

About 74.7% (n = 56) of the participants reported a loss in the family, and 77.3% (n = 58) had a history of trauma or had witnessed a traumatic event in the past. Similar findings were reported by Kovess-Masfety et al. (2021), who found that a large majority of Afghan adults residing in Afghanistan, 86.16%, had either experienced or witnessed at least one traumatic event [20]. In addition, 37.3% (n = 28) attempted suicide in the past. Furthermore, 33.3% (n = 25) of the respondents reported substance abuse. Finally, 82.7% (n = 62) believed in or sought faith healing as a remedy. [Table 3]

**Table 3:** Abuse, substance use, and negative life events characteristics of the respondents.

| Variable | Frequency (n) | Percentage (%) |
| --- | --- | --- |
| **Abuse History\*** |  |  |
| Yes | 45 | 67.2 |
| No | 22 | 32.8 |
| **Abuse Type†** |  |  |
| Emotional | 11 | 24.4 |
| Physical | 16 | 35.6 |
| Both | 18 | 40 |
| **Trauma History** |  |  |
| Yes | 58 | 77.3 |
| No | 17 | 22.7 |
| **Loss in Family** |  |  |
| Yes | 56 | 74.7 |
| No | 19 | 25.3 |
| **Substance Abuse** |  |  |
| Yes | 25 | 33.3 |
| No | 50 | 66.7 |
| **Suicide Attempt** |  |  |
| Yes | 28 | 37.3 |
| No | 47 | 62.7 |
| **Faith Healing** |  |  |
| Yes | 62 | 82.7 |
| No | 13 | 17.3 |
| *Percentages are calculated only for ever-married participants (65 married, 2 divorced). |  |  |
| †Percentages are based on the respondents who reported a history of abuse (n = 45). |  |  |

Regarding medication history, a majority of 42.7% (n = 32) of respondents used SSRIs, 16% (n = 12) had a history of TCA use, 13.3% (n = 10) used Benzodiazepines, 9.3% (n = 7) used mood stabilizers, 6.7% (n = 5) used anti-convulsant medication, while 12% (n = 9) used no medications for psychiatric disorders. [Table 4]

**Table 4:**
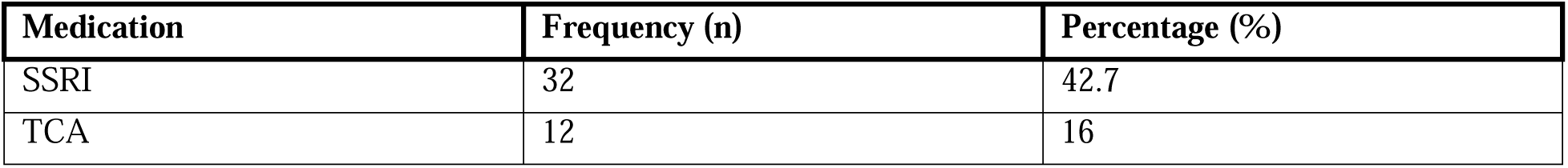
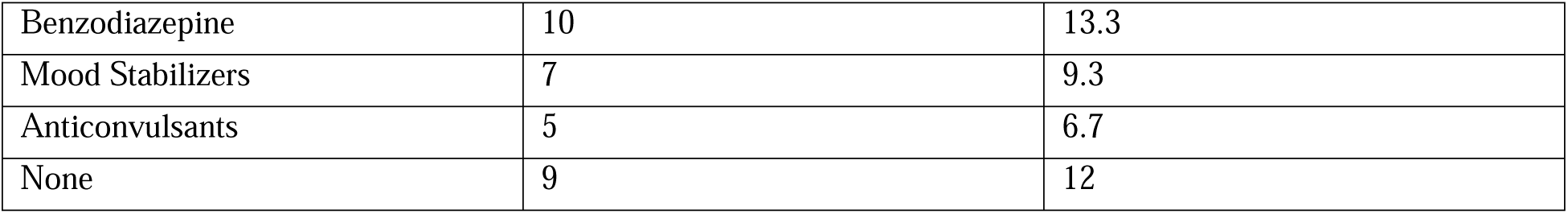
Medication history for psychiatric disorders.

| Medication | Frequency (n) | Percentage (%) |
| --- | --- | --- |
| SSRI | 32 | 42.7 |
| TCA | 12 | 16 |
| Benzodiazepine | 10 | 13.3 |
| Mood Stabilizers | 7 | 9.3 |
| Anticonvulsants | 5 | 6.7 |
| None | 9 | 12 |

A majority of respondents, 62.7% (n = 47), had depression. A total of 33.3% (n = 25) of the participants were without any psychiatric diagnosis, 1.3% (n = 1) had obsessive-compulsive disorder, and 2.7% (n = 2) had bipolar disorder. [Table 5]

**Table 5:** Prevalence of psychiatric diagnoses among study participants.

| Diagnosis | Frequency (n) | Percentage (%) |
| --- | --- | --- |
| Depression (including MDD) | 47 | 62.7 |
| Obsessive-compulsive disorder | 1 | 1.3 |
| Bipolar Disorder | 2 | 2.7 |
| None | 25 | 33.3 |

## DISCUSSION

The current descriptive cross-sectional study is one of the most integrative and inclusive insights into the demographics, psychiatric health, and psychosocial stressors of Afghan refugees residing in Islamabad, Pakistan. This study is inclusive of all genders and surpasses the linguistic barriers that have been a shortcoming in previous studies of this kind. This helps in accurately evaluating the factors that affect the mental well-being of Afghan refugees, such as unemployment, lack autonomy in choosing whom to marry [21], loss of a loved one, domestic violence, illiteracy, lack of sense of security, separation from their community and homeland, survivor’s guilt, and the hardships while fleeing violence.

The findings of this study bring to light the challenging sociodemographic profile and mental health statistics of the Afghan refugee population residing in Pakistan. Within this clinic-based sample, most of the population is uneducated, has undergone arranged monogamous marriages, has 5-8 children, and more than half of the sample is unemployed. There exists a notable inequality in the education status between males and females in our sample population. A concerning proportion of female respondents (64.6%) had not received formal education, while most of the male participants (85%) were educated. This reflects the structural barriers to female education historically present in Afghan society, which, despite sustained reformed initiatives, remain difficult to overcome [22]. In addition, a significant proportion (69.2%) of married refugees in our study reported a history of physical and/or emotional abuse. These findings are comparable to those of a cross-sectional study conducted by Shinwari et al. (2021) among ever-married Afghan women residing in Afghanistan, which reported a high prevalence of physical abuse (50.52%) and emotional violence (37.4%). This relatively higher prevalence among the refugee population observed in our study may be attributable to various factors which include shifts in gender role identities, unstable interpersonal relationships and disrupted household structure and dynamics, all of which potentially lead to higher rates of both emotional and physical abuse as elaborated in the study by Hyder et al. (2007) [23]. Furthermore, the high prevalence of intimate partner violence (IPV) observed in this study can be attributable to the majority of the female respondents in our study being uneducated, given that primary education is a known protective factor against IPV [24]. The observed high prevalence of abuse and female illiteracy align with the broader context of Afghanistan’s patriarchal beliefs, daily stressors, limited infrastructure, restricted access to healthcare and education, lack of family planning, and ongoing conflict [25,26].

Moreover, a noteworthy number (33.3%) of the respondents reported substance abuse, which is markedly higher than the prevalence (5.03%) reported in a study by Sabawoon et al. (2025) regarding substance abuse in the non-refugee general Afghan population [27]. This elevated prevalence in the refugees may be attributable to cumulative stress resulting from both pre-existing vulnerabilities and displacement-related stressors, exacerbating their risk of substance abuse [28]. Furthermore, a concerning 37% of study participants reported previous suicide attempts, emphasizing the necessity for a comprehensive suicide risk assessment for all refugee patients presenting to the OPD, since a prior suicide attempt is one of the major risk factors for future attempts [29]. Finally, a significant number of participants had a documented history of depression and were concurrently prescribed SSRIs, which aligns with the prevalence of SSRI prescriptions for depression among patients attending tertiary care psychiatry outpatient departments, as in our setting [30]. Remarkably, 82.7% of the participants believed in faith healing, which underscores the importance of religiously and culturally sensitive integration of these belief systems for addressing and treating the mental health issues faced by the Afghan population, to improve the quality of health care and maintain inclusivity.

The findings of this study can be added upon by overcoming the bias and conducting a similar descriptive study in other urban and rural health centers of Pakistan. The population selection can also be made more inclusive by conducting this study on a wider scale since there is a huge stigma when it comes to seeking mental help for oneself amongst Afghans, necessitating further research to be conducted in educational institutes, residential communities, and refugee camps to better understand the magnitude of mental health issues of this population [31]. Furthermore, a proper questionnaire should be developed for evaluating the indices and frequency of other psychosocial stressors besides the ones evaluated in this study. For example, the population under study can be sought for a history of childhood illnesses, developmental delay, exploitation of labor, religious or cultural intolerance, political oppression, co-existing medical conditions, financial status, community support, etc. A further improvement would be made by comparing similar parameters of the non-refugee population of Afghanistan, evaluating the available interventions for the existing psychiatric morbidities, and comparing the clinical trials of certain pharmaceutical therapies for these morbidities within the Afghan population via meta-analyses.

These findings may help mental health professionals recognize the substantial burden of psychosocial adversity among Afghan refugees attending psychiatric services, and support more systematic assessment of depression, trauma exposure, suicide risk, substance use, and other clinically relevant conditions. This would not only equip our health system with appropriate intervention strategies for Afghan refugees’ psychiatric illnesses but also inculcate empathy in the medical professionals of Pakistan by sensitizing them to torture-induced PTSD [32]. Moreover, it would increase self-awareness among the refugees about the prevalence of psychiatric ailments and their treatment options. This would alleviate the suffering and challenges faced by the victims of political instability and war in Afghanistan by providing them with a chance to rehabilitate and accept the new phase of their lives in Pakistan.

## LIMITATIONS

A key limitation of this study is its limited depth in evaluating psychiatric diagnoses, as the frequency, duration, severity, and impact on quality of life of these conditions were not comprehensively evaluated. Instead, the analysis relied on binary classifications (presence or absence of the diagnoses), potentially overlooking nuanced variations in clinical presentation and their broader implications, such as differences in symptom trajectories. Additionally, the population was selected using convenience sampling within a single tertiary care psychiatry outpatient department, introducing a geographical bias. This, alongside our relatively small sample size limits the generalizability of our study to all the Afghan refugees residing in the country. Additionally, the data was collected via face-to-face interviewing which may have introduced social desirability bias due to the stigma around substance abuse and mental illnesses, resulting in the potential underreporting of the data.

## CONCLUSION

Despite the limitations, the study provides valuable insights into the psychosocial stressors among Afghan refugees residing in Pakistan along with their psychiatric morbidities and respective pharmacological treatment patterns. It emphasizes the importance of clinical treatment, rehabilitative interventions, and culturally sensitive care to counter psychiatric illnesses. Future research, particularly longitudinal and multi-center studies, is required to investigate other significant psychosocial stressors, encompassing a wider geographical scope that includes both rural and urban populations. Lastly, this study highlights an urgent need for researchers and policymakers worldwide to systematically investigate, report, and rehabilitate their refugee population affected by war and other large-scale adversities, particularly by addressing their unique psychosocial stressors and implementing compassionate and culturally sensitive mental health rehabilitation strategies that foster resilience, rebuild trust, and create a sense of belonging for refugee populations worldwide, reaffirming the enduring strength of the human spirit.

## ACKNOWLEDGEMENTS

We thank the participants for their cooperation and the administration of Shifa International Hospital, Islamabad, for facilitating the psychiatric treatment of Afghan refugees and allowing us to design and conduct this research.

## DECLARATIONS

### ETHICS APPROVAL

This study was conducted as per the 1964 Helsinki Declaration of Human Rights and its later amendments. Approval of research synopsis was obtained from the Institutional Review Board & Ethics Committee (IRB & EC) of Shifa International Hospital (SIH), Islamabad (Reference IRB# 490-21). Data was collected after obtaining fully informed, understood, and voluntary consent from the patients.

### PARTICIPATION CONSENT

All participants were 18 years of age or older and provided verbal informed consent before study participation. This procedure was approved by the ethics committee and documented in accordance with the study protocol approved prospectively by the ethics committee. Interviews were conducted in Pashto or Persian according to each participant’s language preference. Only the aggregate findings from participants’ response are reported, and no directly identifying participant information is included in this manuscript. In addition, the dataset used for analysis was accessible only to authorized members of the research team.

### PARTICIPANT SAFETY

Participants reporting current suicidal ideation, an immediate risk of self-harm, ongoing serious abuse, or another urgent safety concern were referred immediately to the attending psychiatrist for clinical risk assessment and management in accordance with the clinic’s established procedures. Participation in the study did not replace or delay routine clinical assessment or treatment.

### DATA AVAILABILITY

The aggregate data supporting the findings of this study are included in the manuscript and its tables. Participant-level data is not publicly available because of participant confidentiality, the sensitive nature of the collected information, and applicable ethical restrictions.

### CONTRIBUTIONS

Mahrukh Nadeem and Abdul Wahab Yousafzai conceptualized and designed the study. Abdul Wahab Yousafzai recruited study participants and administered questionnaires. Mahrukh Nadeem and Abdul Wahab Yousafzai worked on IRB submission and approval. Mahrukh Nadeem and Ahmed Ali Khan compiled the data, performed the formal analysis and interpreted the results. Mahrukh Nadeem, Ahmed Ali Khan, and Muhammad Hussain Mansoor drafted the original manuscript. Ahmed Ali Khan and Muhammad Hussain Mansoor helped in editing and formatting the manuscript. All authors reviewed and approved the final manuscript for submission.

