## Supplementary Appendix 1 for "Psychosocial Stressors and Psychiatric Morbidity Among Afghan Refugees Attending a Psychiatry Clinic in Islamabad, Pakistan: A Descriptive Cross-Sectional Study"

**Supplementary Appendix 1: QUESTIONNAIRE**

**DEMOGRAPHICS AND CLINICAL CHARACTERISTICS OF AFGHAN PATIENTS**

Name: __________________________________________________Age: _______________

Gender:	Male/ Female/ Other

Education:	Secondary/ Intermediate/ Undergraduate/ Postgraduate/ Diploma

Marital status:	Single/ Married/ Divorced/ Separated/ Widowed

Age at time of marriage: __________________________________________________

Type of marriage: _______________________________________________________

No. of Children: _________________________________________________________

Occupation: ____________________________________________________________

History of abuse:	YES/ NO

*If yes,* Type of abuse (emotional/ physical): ___________________________________

History or witness of any threatening or traumatic situation: YES/ NO

History of substance abuse: YES/ NO	History of suicide attempt: YES/ NO

History of faith healing: YES/ NO

Drug History:	SSRI/ Anti-convulsant/ TCA/ Mood stabilizer/ Benzodiazepine

Psychiatric diagnosis: ___________________________________
