## Supplementary Appendix 2 for "Psychosocial Stressors and Psychiatric Morbidity Among Afghan Refugees Attending a Psychiatry Clinic in Islamabad, Pakistan: A Descriptive Cross-Sectional Study"

**Supplementary Appendix 2: PARTICIPATION CONSENT FORM**

**Investigator:**

“I am inviting you to participate in a **research study**. Involvement in the study is **voluntary**, so you may choose to **participate or not**. I am now going to explain the study to you. Please feel free to ask any **questions** that you may have about the research.

“I am interested in learning more about **psychosocial stressors affecting Afghan patients visiting psychiatry clinic in Islamabad, Pakistan**. You will be asked some personal questions regarding your current and past medical issues. This will take approximately 15 minutes of your time. All information will be kept **confidential***.* If you choose not to mention your name (stay anonymous), this means that **your name will not appear anywhere and no one except me will know about your specific answers**. Your responses will be assigned a code number, and only the researcher will have the key to indicate which number belongs to which participant.

“The benefit of this research is that you will be helping us to understand **psychosocial stressors affecting Afghan patients visiting psychiatry clinic in Islamabad, Pakistan***.* This information should help us to **understand issues affecting the mental health of Afghans seeking treatment here***.* The risks to you for participating in this study are **none***.* These risks will be minimized by ensuring your **confidentiality***.* If you do not wish to continue, you have the right to withdraw from the study at any time.”

**Participant:**

“All of my questions and concerns about this study have been addressed. I choose, voluntarily, to participate in this research project. I certify that I am at least 18 years of age.”

Print name of participant: ________________________________________________________________

Signature of participant: __________________________________	Date: ___________________
